# A Prospective Randomized Crossover Trial of Systemic Chemotherapy in Patients with Low-Grade Mucinous Appendiceal Adenocarcinoma

**DOI:** 10.1101/2022.12.06.22283164

**Authors:** John Paul Shen, Abdelrahman M. Yousef, Fadl A. Zeineddine, Mohammad A. Zeineddine, Rebecca S. Tidwell, Karen A. Beaty, Lisa C. Scofield, Safia Rafeeq, Nick Hornstein, Elizabeth Lano, Cathy Eng, Aurelio Matamoros, Wai Chin Foo, Abhineet Uppal, Christopher Scally, Paul Mansfield, Melissa Taggart, Kanwal P. Raghav, Michael J. Overman, Keith Fournier

## Abstract

**Importance:** Appendiceal Adenocarcinoma is a rare tumor and given the inherent difficulties in performing prospective trials in such a rare disease currently there is a scant amount of high-quality data upon which to guide treatment decisions, which highlights the unmet need for more pre-clinical and clinical investigation for this orphan disease

**Objective:** To objectively evaluate the effectiveness of flouropyrimdine-based systemic chemotherapy in inoperable low-grade mucinous Appendiceal Adenocarcinoma patients.

**Design:** This open label randomized crossover trial recruited patients from September 2013 to January 2021. The data collection cutoff was May 2022.

**Setting:** Single tertiary care comprehensive cancer center.

**Participants:** Enrollment of up to 30 patients was planned. Eligible patients had histological evidence of a metastatic low grade, mucinous Appendiceal Adenocarcinoma, with radiographic images demonstrating the presence of mucinous peritoneal carcinomatosis and were not considered a candidate for complete cytoreductive surgery. Key exclusion criteria were concurrent or recent investigational therapy, evidence of a bowel obstruction, use of total parental nutrition.

**Interventions:** Patients were randomized to either 6 months observation followed by 6 months of chemotherapy, or initial chemotherapy followed by observation. The majority of patients were treated with either 5FU or capecitabine as single agent (n = 15, 63%); 3 (13%) received doublet chemotherapy (FOLFOX or FOLFIRI), bevacizumab was added to cytotoxic chemotherapy for 5 (21%) patients.

**Main Outcomes and Measures:** The difference in tumor growth and patients reported outcomes between the chemotherapy and observation periods. Also, the objective response rate, the rate of bowel complications, and differences in overall survival.

**Results:** A total of 24 patients were enrolled. Fifteen patients were available to evaluate difference in tumor growth between treatment and observation; there was not a significant difference (8.4% (1.5, 15.3%) increase from baseline on treatment vs. 4.0% (−0.1, 8.0%) increase from baseline on observation; p=0.26). Of the 18 patients who received any chemotherapy, zero had an objective response (14 (77.8%) SD, 4 (22.2 %) PD). Median OS was 53.2 months, there was no significant difference in OS between the Observation First arm (76 months) and the Treatment First arm (53 months) (HR, 0.64; 95% CI, 0.16 to 2.6; p = 0.48). Patient reported quality of life metrics identified that fatigue (Mean scores were 18.5 vs 28.9, p=0.02), peripheral neuropathy (6.7 vs 28.9, p=0.014), and financial difficulty (8.9 vs 28.9, p=0.0013) were all significantly worse while on treatment.

**Conclusions and Relevance:** These data suggest that patients with low-grade mucinous appendiceal adenocarcinoma do not derive benefit from systemic fluoropyrimidine-based chemotherapy.

**Trial Registration:** ClinicalTrials.gov Identifier: NCT01946854.

URL: https://clinicaltrials.gov/ct2/show/NCT01946854

**KEY POINTS:** *Question:* Is fluoropyrimidine-based systemic chemotherapy effective in treating inoperable low-grade mucinous Appendiceal Adenocarcinoma patients?

*Findings:* In this randomized clinical trial that included 24 patients, there was no significant difference in tumor growth between treatment and observation (8.4% increase from baseline on treatment vs. 4.0% increase from baseline on observation; p=0.26).

*Meaning:* Patients with low-grade mucinous appendiceal adenocarcinoma do not derive benefit from systemic fluoropyrimidine-based chemotherapy.

## INTRODUCTION

Appendiceal adenocarcinoma (AA) is both a rare and heterogenous disease, with marked contrast in the natural history of low-grade and high-grade tumors (5-year overall survival (OS) 68% for low-grade vs. 7% for high-grade).^1-4^ The rarity of AA has made it difficult to study with traditional prospective, randomized controlled trials; thus, there has been a critical lack of data regarding the responsiveness of appendiceal tumors to chemotherapy. Traditionally, AA has been treated with chemotherapy approved for the treatment of colorectal cancer (CRC) although the evidence to support this practice is primarily anecdotal or in the form of small case series.^5,6^ In the United States, current National Comprehensive Cancer Network (NCCN) guidelines continue to suggest that appendiceal cancer be treated similarly to CRC.^7^ However, low-grade mucinous appendiceal adenocarcinomas represent an extremely unique indolent biology with lack of lymph node and hematogenous spread, limited cytological atypia, and long natural history that are all factors which stand in contrast to CRC.^8-10^ There is also a growing body of molecular data that has identified clear molecular differences between AA and CRC.^2,11-13^ Finally, while literature on chemotherapy response in AA is limited, the few existing reports suggest limited clinical activity of systemic chemotherapy, especially in those patients with mucinous histology and lower grade differentiation.^5,14-18^

Histologically low-grade AA tumors are generally hypocellular with abundant mucin and ‘pushing’, as opposed to infiltrating, margins.^19^ These tumors are known to follow an indolent disease course, and are primarily treated with cytoreductive surgery (CRS) that is often followed by hyperthermic intraperitoneal chemotherapy (HIPEC); this is currently considered the standard-of-care treatment.^14,20-25^ However, despite an absence of prospective data suggesting that low-grade AA patients benefit from systemic chemotherapy, it is common practice that patients with inoperable, low-grade AA are treated with systemic chemotherapy.^18,26-28^ The cytotoxic effects of most traditional chemotherapy drugs such as nucleoside analogs like flouropyrimidine are dependent on the rate of cell division (cell cycle phase specific chemotherapy), which is why it has been hypothesized that the slow growth of this disease would result in intrinsic resistance.^5,29^ Retrospective studies of systemic chemotherapy in low-grade AA have suggested a lack of benefit; these negative results are consistent with our institution’s experience with these low-grade tumors.^14-18^ Therefore, we aimed to conduct the first prospective, randomized crossover trial to objectively evaluate the effectiveness of systemic chemotherapy in low-grade mucinous AA.

In nearly all cases metastatic spread of AA is limited to the peritoneal cavity, causing the clinical syndrome pseudomyxoma peritonei (PMP) in which the peritoneal surfaces and omentum are involved with diffuse gelatinous mucinous implants.^30-32^ Mucinous peritoneal disease is difficult to measure with traditional cross-sectional imaging as it frequently exists as a contiguous erratically shaped area in the peritoneal cavity (**eFigure 1**). In addition, current RECIST criteria do not consider mucinous/cystic disease as measurable. For these reasons, standard RECIST criteria are poorly applicable to AA.^33^ Moreover, AA is a slowly progressive disease and classically-defined thresholds for determining changes in disease extent (typically 20% increase) may take years to occur. Thus, determining systemic chemotherapy benefit through standard outcome measures, such as traditional RECIST response rate and time to disease progression, is not practical in this tumor type. To better quantify peritoneal disease burden mpRECIST, which measures up to 5 areas of mucinous disease in the abdominal cavity, was developed. The general mpRECIST guidelines for tumor evaluation follow the structure established by RECIST version 1.1 with two fundamental differences: (1) up to 5 lesions in the peritoneal cavity are assessed and (2) mucinous lesions are considered measurable disease.

## PATIENTS AND METHODS

### Patients

Eligible patients had histological evidence of a metastatic low grade, defined as well- or well to moderate differentiated, mucinous appendiceal adenocarcinoma, with radiographic images demonstrating the presence of mucinous peritoneal carcinomatosis (PMP) and were not considered a candidate for complete cytoreductive surgery. Surgical candidacy was determined by consultation with Peritoneal Surface Malignancy surgeons at the University of Texas MD Anderson Cancer Center (MDACC) in coordination with our Multidisciplinary Peritoneal Surface Malignancy Conference. Criteria for determining non-resectability were: (1) medical co-comorbidities presenting high surgical risk, (2) tumor bulk and location, such as encasement of the liver hilum or extensive small bowel involvement that would preclude the possibility of obtaining a complete cytoreduction (completeness of cytoreduction score of 0 or 1), or (3) prior cytoreductive surgery that was unsuccessful. Patients were required to have ECOG Performance Status of 0-2, be at least 18 years of age, and have adequate bone marrow function (hemoglobin ≥9.0 g/dl; platelets ≥75,000 cells/mm^3^; absolute neutrophil count ≥1000/mm^3^). Key exclusion criteria were concurrent uncontrolled medical illness that was deemed by the investigator to have the potential to interfere with the delivery of chemotherapy for a six-month time period, concurrent or recent investigational therapy, evidence of a bowel obstruction, use of total parental nutrition, and concurrent non-appendiceal metastatic cancer.

### Study Design and Treatment

Our study was a single-center, open-label, randomized trial with a crossover design. Eligible patients were randomized to one of two arms (**Figure 1**): observation for 6 months followed by chemotherapy for 6 months (Observation First) or chemotherapy for 6 months followed by observation for 6 months (Chemotherapy First). With this crossover design, each patient served as their own control. Randomization was performed through the Computer Randomization Enrollment (CORE) automated telephone randomization system.^34^ While in the chemotherapy condition on each arm, patients were treated with a fluoropyrimidine-based regimen; the specific regimen was selected by the treating physician.

**Fig 1.**
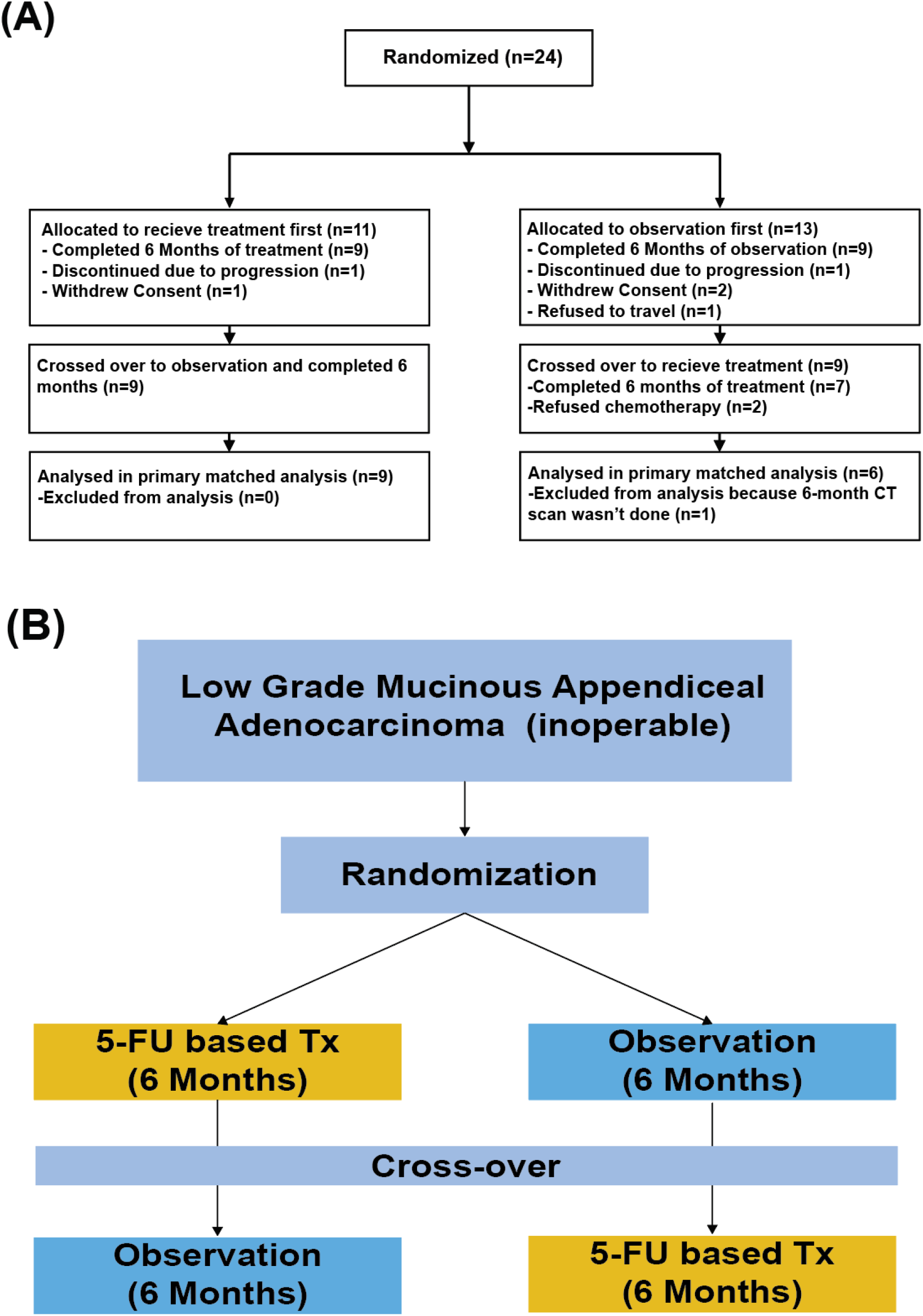
(A) CONSORT flow diagram. (B) Study design. Total study duration is 12 months.

### End points and assessments

The primary end point was the difference in tumor growth (percent change), using the modified peritoneal Response Evaluation Criteria in Solid Tumors (mpRECIST) method, between the chemotherapy and observation periods (regardless of treatment arm). The mpRECIST was chosen due to the known peritoneal only dissemination of this cancer and measured 5 lesions (mucinous and cystic lesions were allowed) in the peritoneal cavity in contrast to the maximum of 2 lesions per standard RECIST. A CT scan of the abdomen and pelvis was performed at baseline and every 3 months as standard of care. Tumor markers (CEA, CA125 and CA 19-9) were measured in peripheral blood collected at baseline, 3, 6, 9 and 12 months. All patients with available 3-, 6-, 9- or 12-month data were combined to compare percent change in each marker level between the observation and treatment periods. Additional secondary efficacy end points were the objective response rate, the rate of bowel complications (defined as bowel obstruction requiring hospitalization or bowel perforation), and differences in OS between early and delayed chemotherapy approaches. Safety monitoring was conducted for the composite safety endpoint of death or bowel complication.

### Patient-Reported Outcomes

An additional secondary end point was difference in quality of life (QoL) between the treatment and observation periods. QoL was determined using three different questionnaires: the European Organization for Research and Treatment of Cancer Core Quality of Life Questionnaire (EORTC QLQ-C30), the ovarian cancer-specific EORTC QoL questionnaire (EORTC QLQ-OV28, due to the considerable similarity in symptomatology of peritoneal disseminated disease from ovarian cancer), and the anxiety-specific Speilberger State/Trait Anxiety Inventory State (STAI) scale.^35,36^ Patients completed the three questionnaires at baseline and every three months.

### Statistical Analysis

To estimate effect size, two readers retrospectively calculated mpRECIST on 5 patients with low grade mucinous AA. The mean change in tumor size over a six-month time period in patients receiving treatments was a 1.6% increase; in those without treatment, the increase was 9%. Based on these preliminary data, a 7.4% effect size (95% Confidence Interval (CI) 3.0%, 11.7%) was observed. The standard deviation of residuals was 3.5% for the random effects introduced by the two readers. Considering both the variation introduced by different readers and variation of the treatment effects, the combined standard deviation of differences was 4.1%. Based on these preliminary data and physician experience, we deemed a ≥5% difference in mpRECIST-determined tumor size change between observation and treatment periods to be clinically meaningful. Assuming a crossover ANOVA square root of mean square error of 4.0% and a one-sided alpha of 0.05, it was estimated that 24 patients would provide 80% power to detect a 5% difference; enrollment of up to 30 patients was planned to have complete 6- and 12-month tumor measures for 24 patients.

Crossover analyses were performed according to Senn’s methods (2002). First, a formal test of interaction and visual inspection for period effect were performed to determine whether the treatment arms (Observation First and Chemotherapy First) could be combined for the test of observation vs. treatment. Subsequently, paired t-tests were used to compare tumor growth after 6 months of observation vs. tumor growth after 6 months of treatment. Patients who did not complete the entire 12-month study period were not included in the primary efficacy analysis. A secondary efficacy analysis including all patients who had any 6-month information was performed using a generalized linear model accounting for the repeated measures for patients with both measures.

A safety monitoring rule was in place to stop the trial early if a Fisher’s exact test ever detected a difference between the treatment and observation period in this composite measure that was ever significant at the 0.05 level. OS was estimated in each arm and graphed by Kaplan-Meier methods. Comparison between the Treatment First and Observation First arms was performed with a log-rank test. Kaplan-Meier curves were implemented in Stata 16 (StataCorp LLC, College Station, TX). All other analyses were performed in SAS 9.4 (The SAS Institute Inc., Cary, NC).

## RESULTS

### Patient Characteristics and Disposition

Between December 2013 and January 2021, a total of 24 patients were randomized to either Chemotherapy First (N=11) or Observation First (13). The median age was 63 years and there was an equal proportion of men and women enrolled; all patients had ECOG performance status of 0 or 1. The majority of patients (n=20, 83%) had well-differentiated tumors, (n=4, 17%) had well-to-moderately differentiated tumors. Pathological diagnosis was confirmed by a pathologist with specific expertise in appendiceal cancer, and graded using a three tiered system evaluating tumor cellularity, destructive invasion, presence of signet ring cells, as well as complexity of tumor architecture^37,38^ (**eTable 1**). Sixteen patients had any tumor mutation testing performed as part of standard-of-care treatment. As expected *KRAS* (n=11, 69%) and *GNAS* (n=8, 57%) were the most frequently mutated genes, there were also two patients with *TP53* mutation (**eTable 2**).^2,8^ None of the tumors had mutation in *APC*. Notably, nearly all of the patients had prior cytoreductive surgery (n= 22, 92%). There was a wide range in the time from diagnosis to randomization with 3 patients (13%) randomized within six months of diagnosis and 6 (25%) randomized over five years from initial diagnosis. After randomization, the Chemotherapy First and Observation First arms were balanced with respect to these categories (**eTable 3**). Due to slow accrual, after eight years of recruitment, the trial was stopped with 24 of the planned 30 patients enrolled. Three patients withdrew consent prior to completing the first 6 month period and were excluded from the primary endpoint analysis (one patient had difficulty scheduling and insurance issues, one patient was unable to maintain follow up because of family issues and one patient couldn’t travel because of the hurricane and being involved with the relief program). Two patients in the Observation First arm completed the observation period but then declined further treatment, and one patient declined traveling to MD Anderson (**Figure 1**). The majority of patients were treated with either 5FU or capecitabine as single agent (n = 15, 63%); bevacizumab was added for three patients (13%), one patient was treated with FOLFOX, and two patients were treated with FOLFIRI (8%).

### Efficacy

Fifteen patients completed the full 12 month study period and were available to evaluate the primary endpoint of difference in tumor growth between treatment and observations periods; there was not a significant difference with trend towards more growth on treatment (8.4% increase on treatment vs. 4.0% increase on observation; p=0.26, **eTable 4**). The interaction between timing and treatment was not statistically significant (p=0.60) in the crossover analysis and minimal upon visual inspection (**eFigure 2**). Therefore, a simple paired analysis of the percent change in tumor volume on treatment vs. observation was conducted for all 18 patients with measurements in both conditions. Similarly, rate of tumor growth was not significantly different between treatment and observation periods (13.1% increase on treatment vs. to 4.4% increase on observation; p=0.37) with trend toward more growth on treatment (**Figure 2**). In total 18 patients received chemotherapy during the study period and zero achieved an objective response, 14 (77.8%) had stable disease during the entire year of follow up, 4 (12.2%) had progression on study (**Figure 2D**).

**Fig 2.**
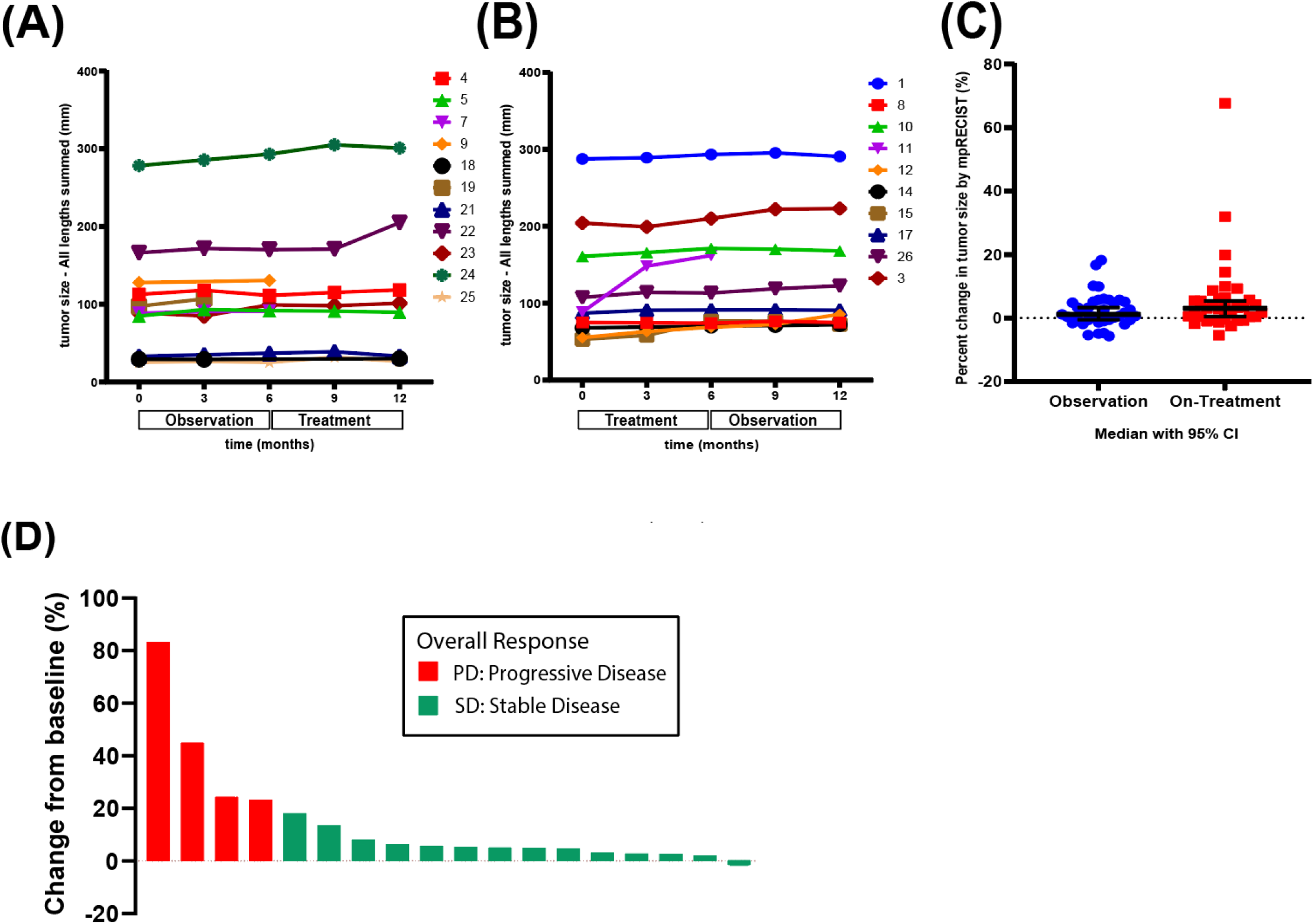
Spider plots showing tumor growth over time in observation first arm (A) and tumor growth overtime in treatment first arm (B). (C) Percent change in tumor size by mpRECIST method between observation and treatment. A novel quantitative measuring system designed for mucinous peritoneal disease, measures up to 5 areas of mucinous disease in the abdominal cavity. (D) Waterfall plot showing best overall response after treatment period

Median OS for the entire cohort was 53.2 months, there was no significant difference in OS between the Observation First arm (76 months) and the Treatment First arm (53 months) (HR, 0.64; 95% CI, 0.16 to 2.6; p = 0.48). The median duration of follow up after study completion at the time of data cutoff was 27 months (8 to 95 months), only three (14.3%) patients received further systemic treatment after the trial (**Figure 3**). There was no significant difference between observation and treatment periods for the percent change in any of the tumor markers evaluated, CEA, CA-125 and CA 19-9; (**eFigure 3)**. The mean percent change in tumor markers for observation and treatment were 50% vs 4% for CEA (p=0.23), 2% vs 2% for CA-125 (p=0.99), and 13% vs 6% for CA 19-9 (p=0.39). Notably the two patients with greatest elevation in CEA and CA 125 (patients 3 and 7, respectively; **Figure 4**); had markedly worse outcome with 22 and 10-month OS.

**Fig 3.**
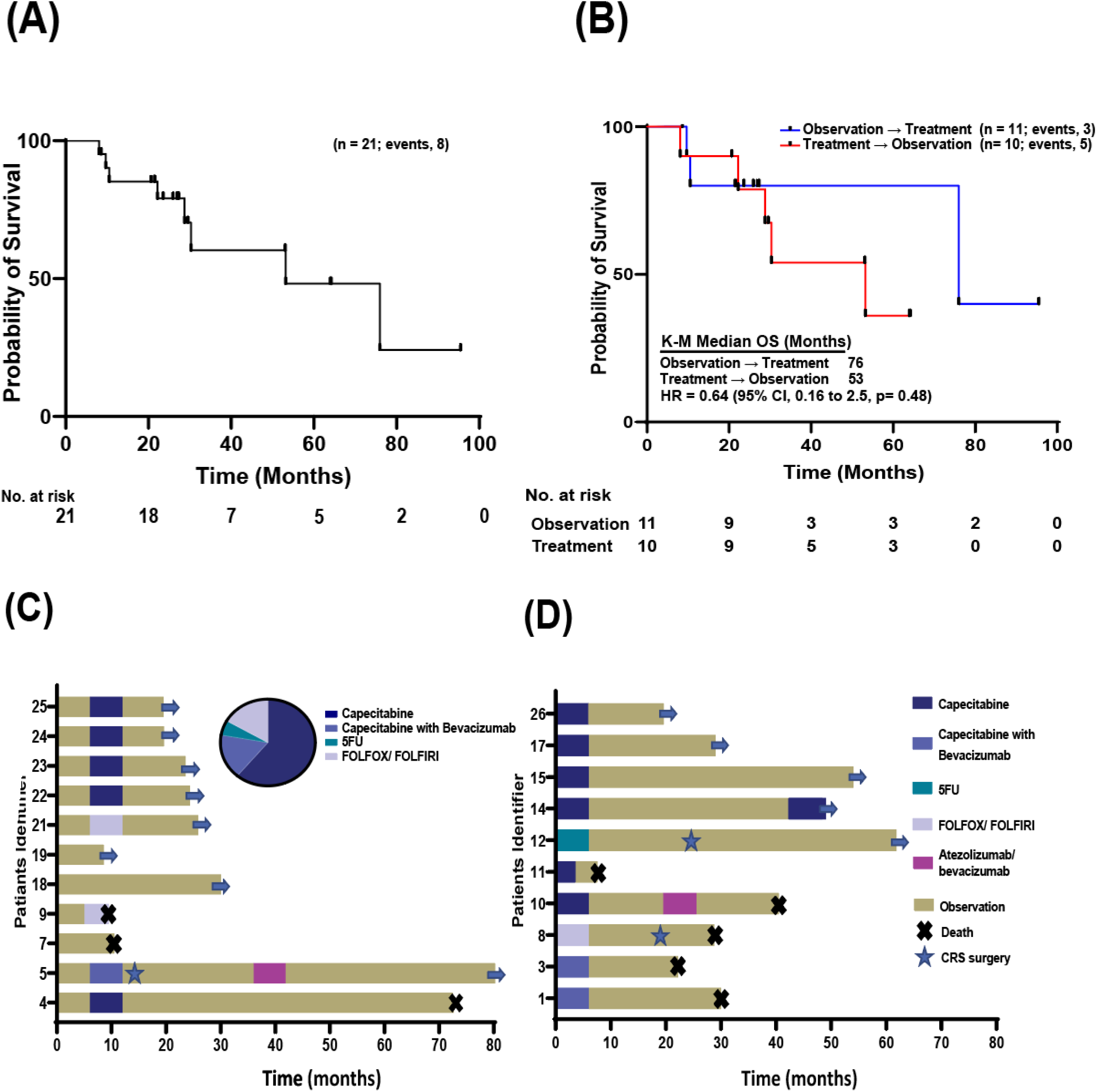
Kaplan Meier curves showing Overall survival of all patients (A) and between the two groups (B). Swimmer plots showing treatment history overtime for observation first arm (C) and Treatment first arm (D). The pie chart shows the chemotherapy distribution patients received during the trial period

**Fig 4.**
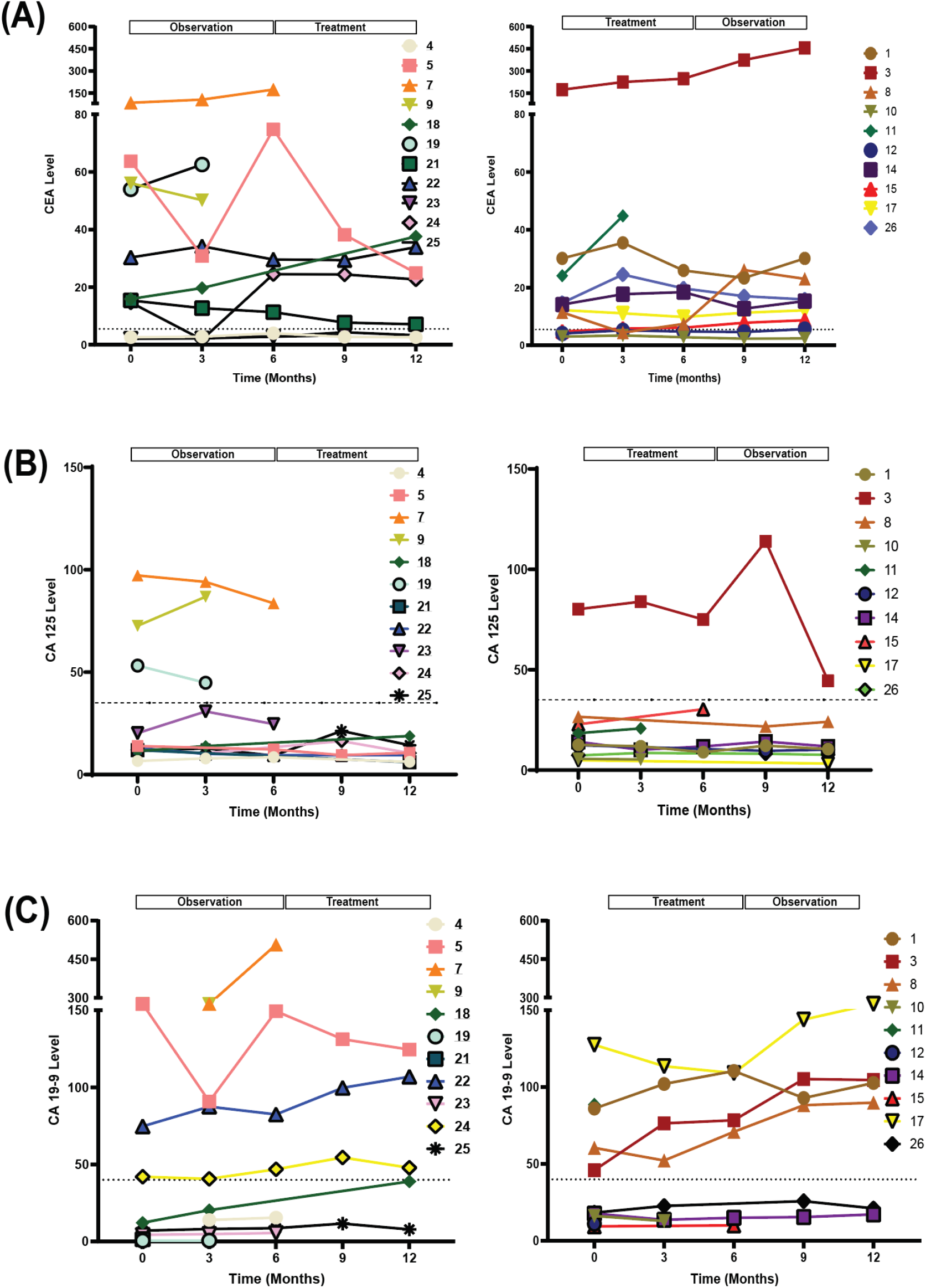
Spider plots showing tumor markers level over time. (A) CEA levels in observation-first arm on the left and treatment-first arm on the right. (B) CA 125 levels in observation-first arm on the left and treatment-first arm on the right. (C) CA19-9 levels in observation-first arm on the left and treatment-first arm on the right.

### Safety

The composite safety outcome measure was similar between treatment and observation and between the 2 arms (**eTable 5)**. Three patients died while on trial, two during the treatment period and one during the observation period. No patients had a bowel perforation during treatment or observation. Four patients were hospitalized for bowel obstruction, two each from Treatment First and Observation First cohorts. Of note one of these patients had four separate admissions (twice during treatment and twice during observation) and was counted once during each time period in eTable 5.

### Patient-Reported Outcomes

Fifteen patients completed the patient-reported outcome questionnaires at both 6 and 12 months and were available for paired analyses. EORTC QLQ-C30 role function score (RF), fatigue score (FA) and financial difficulties (FI) scores were significantly increased during treatment relative to observation indicating worse quality of life while receiving chemotherapy (**Figure 5**); mean scores following observation versus following treatment, respectively, were 92 vs 82 for RF (p=0.03); 19 versus 29 for FA (p=0.02) and 9 versus 29 for FI (p=0.01) (**eTable 6**). EORTC QLQ-OV28 scores for peripheral neuropathy (PN, 6.67 vs 39.89, p=0.001) and chemotherapy side effect (CH, 16.45 vs 23.11, p=0.008) were significantly higher while on treatment (**eTable 7**). There was no significant difference in State Anxiety or Trait Anxiety scores between observation and treatment periods (**eTable 8**).

**Fig 5.**
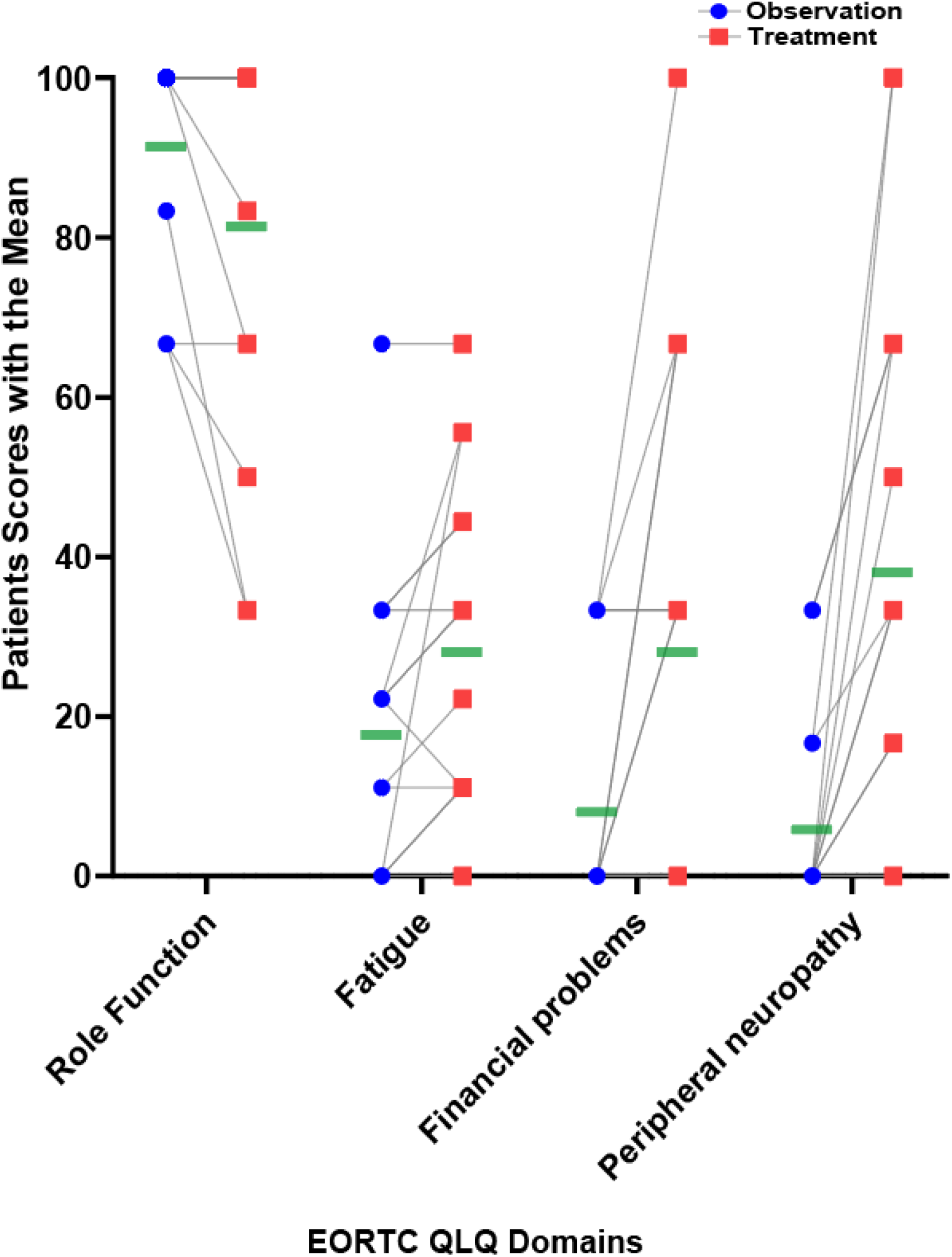
Quality of Life Questionnaire (EORTC QLQ) individual scores between observation and treatment. Green bar represents mean score

## DISCUSSION

Due both to the rarity and heterogeneity of AA, it has been difficult to objectively determine if systemic chemotherapy is effective in the treatment of this disease. This study represents the first prospective, randomized, trial for low-grade mucinous AA to answer this pivotal question. Based on these data fluoropyrimidine-based chemotherapy appears to be ineffective for patients with low-grade mucinous AA, as there was no significant difference in tumor growth, or tumor markers, between the observation vs. treatment periods. Moreover, chemotherapy significantly decreased QoL during the chemotherapy period relative to the observation period. Similarly, delaying the start of chemotherapy with a six-month observation period did not reduce OS nor increase rate of bowel perforation or obstruction, the most feared consequences of progressive peritoneal carcinomatosis.

The results from this prospective, randomized, trial are consistent with multiple prior retrospective analyses suggesting chemotherapy is ineffective in low-grade AA.^14,15,17,18,28,39^ *Shaib et al* found that patients with metastatic low-grade appendiceal mucinous neoplasms who did not receive systemic chemotherapy had longer median OS than those who did (82 vs 32 months, p = .044).^16^ Similarly, a retrospective study that included 1919 metastatic low-grade mucinous AA patients from the National Cancer Database (NCDB) from 1985-2006 found chemotherapy was not associated with improved survival histology (HR 0.95; 95% CI, 0.86-1.04, p = 0.3).^15^ A more recent analysis of NCDB data from 2004-2015 including 639 metastatic low-grade mucinous AA patients confirmed this lack of survival benefit (HR 1.1, 95% CI: 0.82-1.4, p = 0.6).^18^ With respect to perioperative systemic chemotherapy, in a retrospective analysis of 104 patients with PMP of mixed grades who underwent CRS/HIPEC in a multivariate analysis including grade, preoperative chemotherapy was associated with worse survival (HR 2.7, p = 0.03).^14^ Similarly, a retrospective study of perioperative chemotherapy in 284 mucinous PMP patients found that for the 22 low-grade patients treated with systemic chemotherapy there was no difference in either OS or PFS relative to a matched cohort without chemotherapy (PFS 29.5 months with chemotherapy vs. 37.0 months with CRS/HIPEC alone, p = 0.18). ^17^

To our knowledge there are no reports of objective response from cytotoxic systemic chemotherapy specifically in low grade AA. It is important to note that this is in contrast to high-grade AA which is known to be responsive to cytotoxic chemotherapy on the basis of many prior reports^28,40^. Of note there is only one other published prospective trial of systemic chemotherapy in unresectable PMP, a single arm Phase II study of mitomycin C and capecitabine which showed tumor reduction in 6 (15%), stable disease in 18 (45%) and progression in 11 (27.5%) of patients^41^. However, this study included a mixed population of tumors with 32% (13 of 40 patients) higher grade MAA classified as mucinous carcinoma peritonei (PMCA) and 68% (17 or 40 patients) low-grade MAA classified as disseminated peritoneal adenomucinous (DPAM)^42,43^. This study did not breakdown the response by histology, so it is unclear if any of the six responding patients had low-grade tumors. Although several retrospective studies that combined both high- and low-grade AA have reported an aggregate benefit to chemotherapy^5,41,44^, the results of this prospective study and growing recognition of the molecular and clinical differences between high- and low-grade AA^8,45^ argue that these two distinct subtypes should not be grouped together.^46^

We recognize limits to our Phase II study design; the trial was initially planned for enrollment of 30 patients to have complete data for 24 patients, providing 80% power for primary endpoint. However, accrual in this rare disease was slow with only 24 patients enrolled after eight years (2013 to 2021) and was closed due to slow accrual. When designed in 2012 there was concern that delaying start of treatment would harm patients, thus the only pre-specified interim analyses concerned increased death or sever complication in the Observation First arm. Although not pre-specified as trial was not blinded interim efficacy analysis was performed after eight years, given complete lack of response to chemotherapy and no difference in tumor growth between observation and treatment periods the trial was closed as it was felt unethical to continuing treating low-grade AA patients with FU-based chemotherapy. The novel mpRECIST method was determined by the treating physicians during the study period only; thus, there was not consistent way to measure progression after the trial period. All of the patients on study were enrolled at a tertiary referral cancer center, which may not be representative of patients in a community oncology practice. Despite these limitations, our study represents the first prospective, randomized, crossover trial for low-grade AA.

Taking into consideration the lack-of-benefit from fluoropyrimidine-based chemotherapy seen in this trial and prior retrospective studies with similar conclusion these data argue strongly that fluoropyrimidine-based chemotherapy should not be considered the standard-of-care treatment of low-grade AA that are not candidates for CRS. Clinical trials investigating novel therapeutics should be strongly considered for these patients who desire non-surgical anti-cancer therapy. This prospective, randomized clinical data highlight the differences between AA and CRC and demonstrates the need for the development of appendiceal cancer specific guidelines. An additional important observation for the clinical management of low-grade AA is that given natural slow growth of these tumors and the difficulty of imaging peritoneal carcinomatosis, stable disease as assessed by CT or MRI cannot be interpreted as clinical benefit in low-grade AA as it is in most other solid tumors.

In summary, these data from a prospective, randomized crossover design trial indicate that patients with low-grade mucinous AA do not derive benefit from flouropyrimidine-based chemotherapy. We therefore conclude that fluoropyrimidine-based chemotherapy should not be used in this specific patient population. These data demonstrate the unique biology of low-grade mucinous AAs and highlight the need for more pre-clinical and clinical investigation for this orphan disease. ^47-57^

## Data Availability

All data produced in the present work are contained in the manuscript

## Acknowledgements

This work was supported by Golfers Against Cancer, the Col. Daniel Connelly Memorial Fund, the National Cancer Institute (K22 CA234406 to J.P.S., and the Cancer Center Support Grant (P30 CA016672), the Cancer Prevention & Research Institute of Texas (RR180035 to J.P.S., J.P.S. is a CPRIT Scholar in Cancer Research), and a Conquer Cancer Career Development Award to J.P.S. Any opinions, findings, and conclusions expressed in this material are those of the author(s) and do not necessarily reflect those of the American Society of Clinical Oncology® or Conquer Cancer. Writing support provided by Jennifer Peterson, PhD.

---

**eFig 1.**
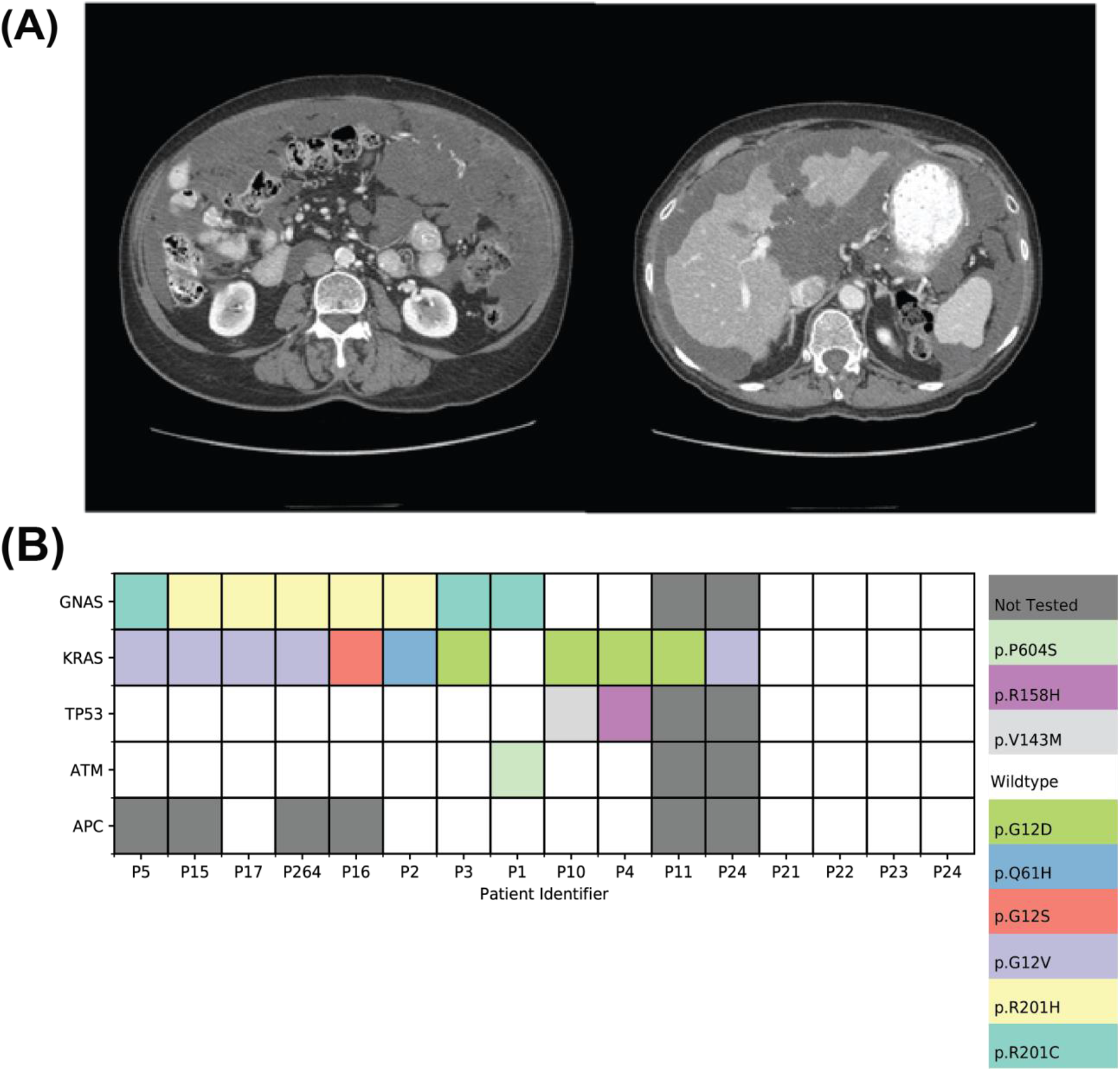
Low-Grade Mucinous Appendiceal Adenocarcinoma. (A) CT scans highlighting diffuse mucinous nature of this tumor type. (B) 16 patients on study has standard-of-care mutation testing, dark grey indicates gene was not tested for that patient, white indicates gene was tested and found to be wildtype.

**eFig 2.**
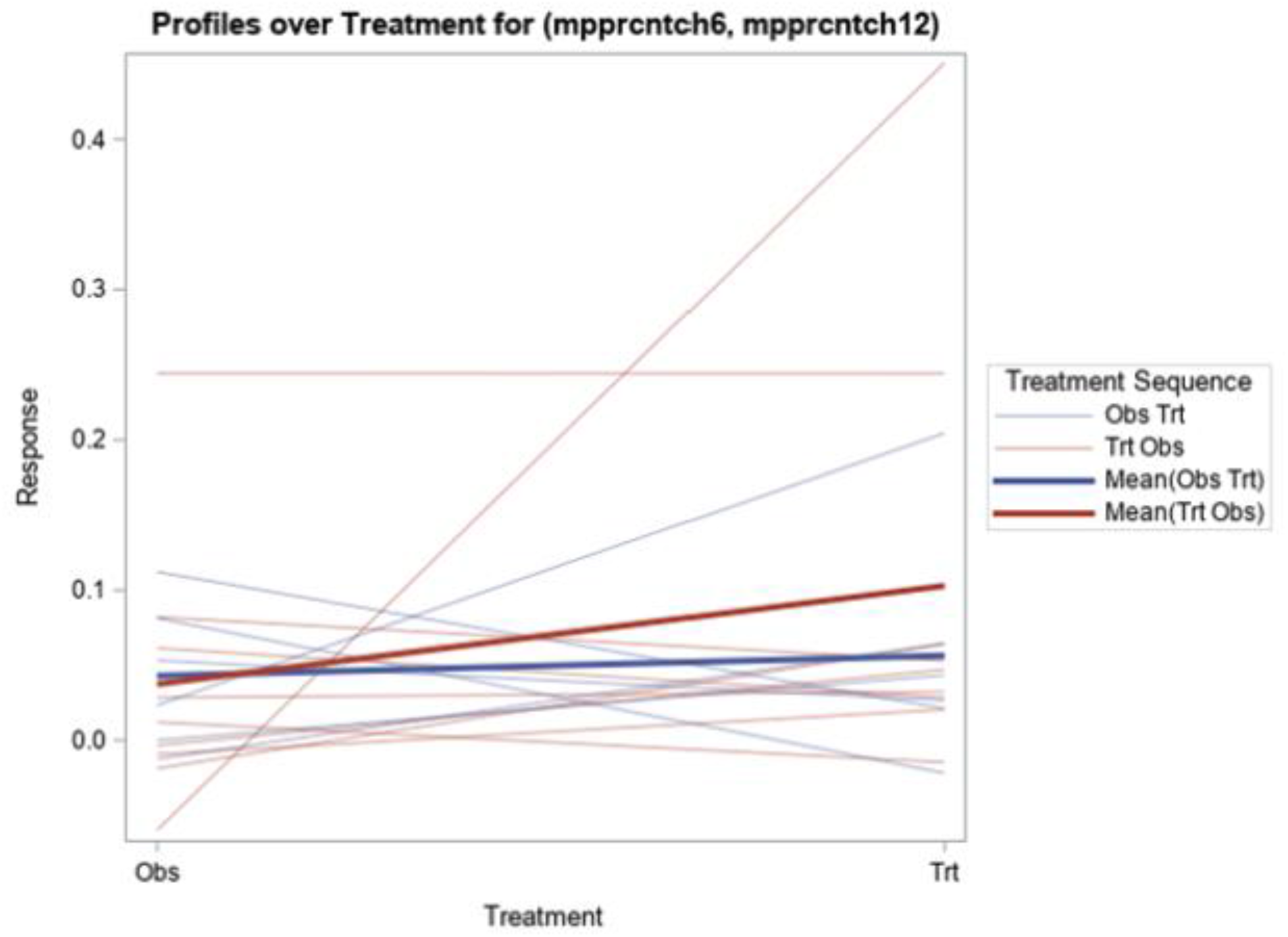
Interaction between time and treatment.

**eFig 3.**
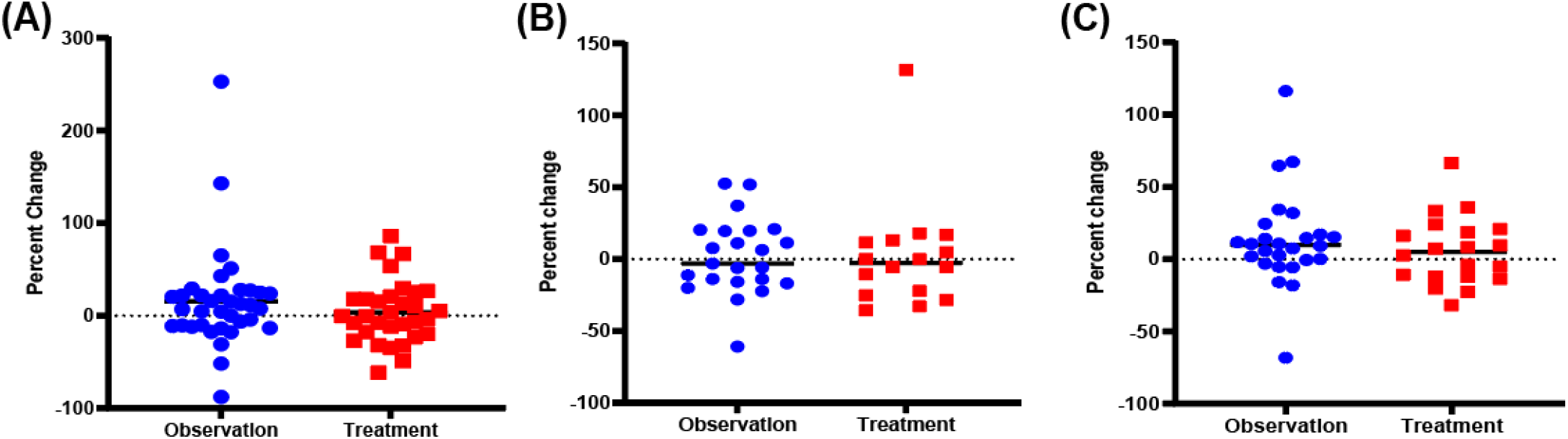
Waterfall plots showing tumor markers percent change between Observation and treatment periods. (A) CEA percent change in observation on the left and treatment on the right. (B) CA 125 percent change in observation on the left and treatment on the right. (C) CA19-9 percent change in observation on the left and treatment on the right.

**eTable 1.** Patients histopathology and grade

| Patient Identifier | Histology | Grade |
| --- | --- | --- |
| 1 | MUCINOUS ADENOCARCINOMA | WELL-TO-MODERATELY DIFFERENTIATED |
| 2 | MUCINOUS ADENOCARCINOMA | WELL-DIFFERENTIATED |
| 3 | MUCINOUS ADENOCARCINOMA | WELL-DIFFERENTIATED |
| 4 | MUCINOUS ADENOCARCINOMA | WELL-TO-MODERATELY DIFFERENTIATED |
| 5 | MUCINOUS ADENOCARCINOMA | WELL-DIFFERENTIATED |
| 6 | MUCINOUS ADENOCARCINOMA | WELL-DIFFERENTIATED |
| 7 | MUCINOUS ADENOCARCINOMA | WELL-DIFFERENTIATED |
| 8 | MUCINOUS ADENOCARCINOMA | WELL-DIFFERENTIATED |
| 9 | MUCINOUS ADENOCARCINOMA | WELL-DIFFERENTIATED |
| 10 | MUCINOUS ADENOCARCINOMA | WELL-DIFFERENTIATED |
| 11 | MUCINOUS ADENOCARCINOMA | WELL-DIFFERENTIATED |
| 12 | MUCINOUS ADENOCARCINOMA | WELL-DIFFERENTIATED |
| 14 | MUCINOUS ADENOCARCINOMA | WELL-DIFFERENTIATED |
| 15 | MUCINOUS ADENOCARCINOMA | WELL-DIFFERENTIATED |
| 16 | MUCINOUS ADENOCARCINOMA | WELL-DIFFERENTIATED |
| 17 | MUCINOUS ADENOCARCINOMA | WELL-DIFFERENTIATED |
| 18 | MUCINOUS ADENOCARCINOMA | WELL-TO-MODERATELY DIFFERENTIATED |
| 19 | MUCINOUS ADENOCARCINOMA | WELL-DIFFERENTIATED |
| 21 | MUCINOUS ADENOCARCINOMA | WELL-DIFFERENTIATED |
| 22 | MUCINOUS ADENOCARCINOMA | WELL-DIFFERENTIATED |
| 23 | MUCINOUS ADENOCARCINOMA | WELL-DIFFERENTIATED |
| 24 | MUCINOUS ADENOCARCINOMA | WELL-DIFFERENTIATED |
| 25 | MUCINOUS ADENOCARCINOMA | WELL-DIFFERENTIATED |
| 26 | MUCINOUS ADENOCARCINOMA | WELL-TO-MODERATELY DIFFERENTIATED |

**eTable 2.** Mutational analysis

| Patient Identifier | Allocation | Genes |  |  |  |  |  |
| --- | --- | --- | --- | --- | --- | --- | --- |
|  |  | GNAS | KRAS | JAK3 | TP53 | ATM | APC |
| 4 | Observation First | Wildtype | p.G12D | Wildtype | p.R158H | wildtype | wildtype |
| 5 | Observation First | p.R201C | p.G12V | wildtype | wildtype | wildtype | N/A |
| 16 | Observation First | p.R201H | p.G12S | wildtype | wildtype | wildtype | N/A |
| 18 | Observation First | p.R201H | p.G12V | wildtype | wildtype | wildtype | N/A |
| 21 | Observation First | wildtype | wildtype | wildtype | wildtype | wildtype | wildtype |
| 22 | Observation First | wildtype | wildtype | p.V617F | wildtype | wildtype | wildtype |
| 23 | Observation First | wildtype | wildtype | wildtype | wildtype | wildtype | wildtype |
| 24 | Observation First | N/A | p.G12V | N/A | N/A | N/A | N/A |
| 1 | Treatment First | p.R201C | wildtype | wildtype | wildtype | p.P604S | wildtype |
| 2 | Treatment First | p.R201H | p.Q61H | wildtype | wildtype | wildtype | wildtype |
| 3 | Treatment First | p.R201C | p.G12D | wildtype | wildtype | wildtype | wildtype |
| 10 | Treatment First | Wildtype | p.G12D | wildtype | p.V143M | wildtype | wildtype |
| 11 | Treatment First | N/A | p.G12D | N/A | N/A | N/A | N/A |
| 14 | Treatment First | wildtype | wildtype | wildtype | wildtype | wildtype | wildtype |
| 15 | Treatment First | p.R201H | p.G12V | wildtype | wildtype | wildtype | N/A |
| 26 | Treatment First | p.R201H | p.G12V | wildtype | wildtype | wildtype | wildtype |

**eTable 3.** Patients characteristics by randomized treatment arm

| Patient Characteristics |  | All<br>N (%) | Treatment First<br>N (%) | Observation First<br>N (%) |
| --- | --- | --- | --- | --- |
| **All** |  | **24 (100%)** | **11 (100%)** | **13 (100%)** |
| Age - median (min, max) | N=24 | 62.8 (38.0, 81.8) | 62.8 (44.8, 79.3) | 60.9 (38.0, 81.8) |
| Race/ethnicity |  |  |  |  |
|  | White or Caucasian | 14 (58%) | 7 (64%) | 7 (54%) |
|  | Black or African | 6 (25%) | 2 (18%) | 4 (31%) |
|  | Any race/ Hispanic | 2 (8%) | 1 (9%) | 1 (8%) |
|  | Other | 2 (8%) | 1 (9%) | 1 (8%) |
| Sex |  |  |  |  |
|  | Female | 12 (50%) | 6 (55%) | 6 (46%) |
|  | Male | 12 (50%) | 5 (45%) | 7 (54%) |
| ECOG PS |  |  |  |  |
|  | 0 | 15 (63%) | 8 (73%) | 7 (54%) |
|  | 1 | 9 (38%) | 3 (27%) | 6 (46%) |
| Histology grade |  |  |  |  |
|  | Well to Moderate differentiated | 4 (17%) | 2 (18%) | 2 (15%) |
|  | Well differentiated | 20 (83%) | 9 (82%) | 11 (85%) |
| Planned treatment |  |  |  |  |
|  | . | 4 (17%) | 0 (0%) | 4 (31%) |
|  | 5FU with Bevacizumab | 1 (4%) | 0 (0%) | 1 (8%) |
|  | 5FU without Bevacizumab | 1 (4%) | 1 (9%) | 0 (0%) |
|  | CAPOX with Bevacizumab | 2 (8%) | 1 (9%) | 1 (8%) |
|  | Capecitabine with Bevacizumab | 2 (8%) | 2 (18%) | 0 (0%) |
|  | Capecitabine without Bevacizumab | 11 (46%) | 6 (55%) | 5 (38%) |
|  | FOLFIRI without Bevacizumab | 2 (8%) | 0 (0%) | 2 (15%) |
|  | FOLFOX without Bevacizumab | 1 (4%) | 1 (9%) | 0 (0%) |
| Observation period complete |  |  |  |  |
|  | No | 6 (25%) | 2 (18%) | 4 (31%) |
|  | Yes | 18 (75%) | 9 (82%) | 9 (69%) |
| Treatment period complete |  |  |  |  |
|  | No | 8 (33%) | 2 (18%) | 6 (46%) |
|  | Yes | 16 (67%) | 9 (82%) | 7 (54%) |
| Prior CRS |  | 22 (92%) | 11 (100%) | 11 (85%) |
| Time from diagnosis to randomization (months) |  |  |  |  |
|  | 0 to 6 | 3 (13%) | 1 (9%) | 2 (15%) |
|  | 7 to 23 | 9 (37%) | 3 (27%) | 6 (47%) |
|  | 24 to 60 | 6 (25%) | 3 (27%) | 3 (23%) |
|  | >60 | 6 (25%) | 4 (37%) | 2 (15%) |
| KRAS and GNAS status |  |  |  |  |
|  | N = 16 | 16 (100%) | 8 (100%) | 8 (100%) |
|  | KRAS Mutated | 11 (69%) | 6 (75%) | 5 (63%) |
|  | GNAS Mutated | 8 (50%) | 5 (63%) | 3 (38%) |

PF = Performance Status 5FU = 5 Fluorouracil CRS = Cytoreductive Surgery

**eTable 4.**
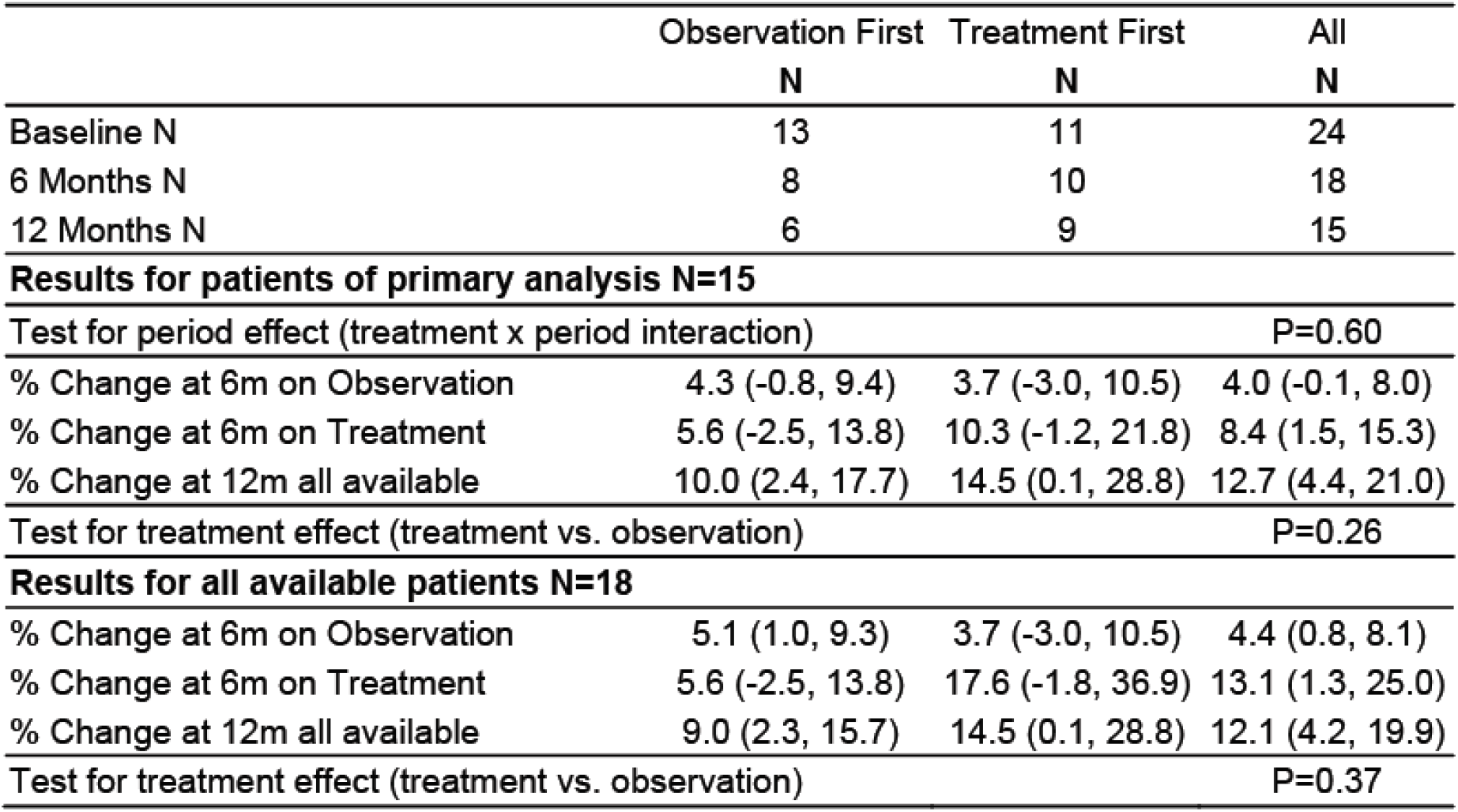
Tumor Measure Availability and Percent Change for Evaluable Patients

**eTable 5.** Numbers of patients with Monitored Adverse Events, counted once per treatment period

| Toxicity |  | Treatment 1<sup>st</sup> | Observation 1<sup>st</sup> | Total |
| --- | --- | --- | --- | --- |
| **N Evaluable\*** |  | **10** | **11** | **21** |
| **Bowel Obstruction\*\*** | Observation | 1 | 1 | 2 |
|  | Treatment | 2 | 1 | 3 |
| **Bowel Perforation** | Observation | 0 | 0 | 0 |
|  | Treatment | 0 | 0 | 0 |
| **Deaths\*\*\*** | Observation | 1 | 0 | 1 |
|  | Treatment | 0 | 2 | 2 |
| **Any Events** | Observation | 1 | 1 | 2 |
|  | Treatment | 2 | 1 | 3 |

\* To be evaluable for toxicity monitoring, the patient either needed to have had an event or completed 3 months on study with no toxic event
\*\* Obstruction counted only when it required hospitalization. The table numbers include one patient on the treatment first arm who had bowel obstruction hospitalizations twice each on both treatment and observation, for a total of 8 obstruction events among 4 patients
\*\*\* Deaths counted only while the patient was actively on study treatment or observation

**eTable 6.** Paired t-test for QLQ C30

|  | P value | Mean of Observation | Mean of Treatment | Difference | SE of difference | t ratio | df |
| --- | --- | --- | --- | --- | --- | --- | --- |
| Global health status/QoL | 0.770030 | 75.00 | 73.33 | 1.665 | 5.587 | 0.2981 | 14.00 |
| Physical Function | 0.070271 | 90.11 | 84.00 | 6.111 | 3.119 | 1.960 | 14.00 |
| **Role Function** | **0.033462** | **92.22** | **82.22** | **10.00** | **4.242** | **2.358** | **14.00** |
| Emotional Function | 0.653262 | 86.67 | 88.89 | -2.223 | 4.844 | 0.4590 | 14.00 |
| Cognitive Function | 0.684669 | 83.33 | 85.56 | -2.223 | 5.362 | 0.4147 | 14.00 |
| Social Function | 0.138278 | 92.22 | 85.56 | 6.667 | 4.241 | 1.572 | 14.00 |
| **Fatigue** | **0.024771** | **18.52** | **28.89** | **-10.37** | **4.125** | **2.514** | **14.00** |
| Nausea / vomiting | 0.510225 | 10.00 | 6.667 | 3.333 | 4.933 | 0.6757 | 14.00 |
| Pain | 0.817841 | 14.44 | 15.56 | -1.111 | 4.732 | 0.2347 | 14.00 |
| Dyspnoea | 0.189362 | 14.29 | 21.43 | -7.143 | 5.157 | 1.385 | 13.00 |
| Insomnia | 0.806295 | 22.22 | 24.44 | -2.221 | 8.889 | 0.2499 | 14.00 |
| Appetite loss | 0.271142 | 4.444 | 11.11 | -6.667 | 5.819 | 1.146 | 14.00 |
| Constipation | 0.272192 | 11.90 | 19.05 | -7.144 | 6.230 | 1.147 | 13.00 |
| Diarrhoea | 0.510316 | 15.56 | 22.22 | -6.666 | 9.867 | 0.6756 | 14.00 |
| **Financial difficulties** | **0.014008** | **8.888** | **28.89** | **-20.00** | **7.127** | **2.806** | **14.00** |

**eTable 7.** Paired t-test for QLQ OV28

|  | P value | Mean of Observation | Mean of Treatment | Difference | SE of difference | t ratio | df |
| --- | --- | --- | --- | --- | --- | --- | --- |
| Abdominal / GI | 0.985919 | 19.26 | 19.33 | -0.07533 | 4.193 | 0.01797 | 14.00 |
| **Peripheral neuropathy** | **0.001303** | **6.667** | **38.89** | **-32.22** | **8.045** | **4.005** | **14.00** |
| Hormonal | 0.999917 | 14.44 | 14.44 | 0.0006667 | 6.300 | 0.0001058 | 14.00 |
| Body image | 0.334376 | 15.56 | 11.11 | 4.443 | 4.444 | 0.9998 | 14.00 |
| Attitude to disease/ treatment | 0.344322 | 23.70 | 28.89 | -5.185 | 5.297 | 0.9787 | 14.00 |
| **Chemotherapy side effects** | **0.007997** | **16.45** | **23.11** | **-6.667** | **2.158** | **3.090** | **14.00** |
| Other single items | 0.081485 | 5.185 | 13.15 | -7.963 | 4.242 | 1.877 | 14.00 |

**eTable 8.** Paired t-test for STAI

|  | P value | Mean of Observation | Mean of Treatment | Difference | SE of difference | t ratio | df |
| --- | --- | --- | --- | --- | --- | --- | --- |
| State Anxiety | 0.217648 | 50.71 | 49.74 | 0.9613 | 0.7447 | 1.291 | 14.00 |
| Trait Anxiety | 0.067773 | 52.57 | 51.22 | 1.347 | 0.6807 | 1.979 | 14.00 |

